# Classification of glomerular pathological findings using deep learning and nephrologist–AI collective intelligence approach

**DOI:** 10.1101/2019.12.30.19016162

**Authors:** Eiichiro Uchino, Kanata Suzuki, Noriaki Sato, Ryosuke Kojima, Yoshinori Tamada, Shusuke Hiragi, Hideki Yokoi, Nobuhiro Yugami, Sachiko Minamiguchi, Hironori Haga, Motoko Yanagita, Yasushi Okuno

## Abstract

**Background:** Automated classification of glomerular pathological findings is potentially beneficial in establishing an efficient and objective diagnosis in renal pathology. While previous studies have verified the artificial intelligence (AI) models for the classification of global sclerosis and glomerular cell proliferation, there are several other glomerular pathological findings required for diagnosis, and the comprehensive models for the classification of these major findings have not yet been reported. Whether the cooperation between these AI models and clinicians improves diagnostic performance also remains unknown. Here, we developed AI models to classify glomerular images for major findings required for pathological diagnosis and investigated whether those models could improve the diagnostic performance of nephrologists.

**Methods:** We used a dataset of 283 kidney biopsy cases comprising 15888 glomerular images that were annotated by a total of 25 nephrologists. AI models to classify seven pathological findings: global sclerosis, segmental sclerosis, endocapillary proliferation, mesangial matrix accumulation, mesangial cell proliferation, crescent, and basement membrane structural changes, were constructed using deep learning by fine-tuning of InceptionV3 convolutional neural network. Subsequently, we compared the agreement to truth labels between majority decision among nephrologists with or without the AI model as a voter.

**Results:** Our model for global sclerosis showed high performance (area under the curve: periodic acid-Schiff, 0.986; periodic acid methenamine silver, 0.983); the models for the other findings also showed performance close to those of nephrologists. By adding the AI model output to majority decision among nephrologists, the sensitivity and specificity were significantly improved in 9 of 14 constructed models compared to those of nephrologists alone.

**Conclusion:** Our study showed a proof-of-concept for the classification of multiple glomerular findings in a comprehensive method of deep learning and suggested its potential effectiveness in improving diagnostic accuracy of clinicians.

## 1. Introduction

Renal pathology is important for the diagnosis and management of patients with kidney disease. The renal survival rate tends to be better with histologic evaluation by renal biopsy than without renal biopsy [1], thus, accurate and robust diagnosis is essential for the proper management of patients with kidney disease. On the other hands, making an accurate diagnosis is a time-consuming process even for experienced pathologists. It has been expected that automated processing to support this procedure will improve the efficiency of renal pathology and contribute to a more objective and standardized diagnosis [2], especially in hospitals, areas, or countries where there are an insufficient number of nephropathologists. A field called digital pathology, which aims to diagnose and quantify disease based on image data obtained by scanning pathological tissue specimens, has rapidly been developed. With the use of current state-of-the-art techniques of deep learning (DL), the artificial intelligence (AI) approach has made a significant progress in medical image analysis of retinal fundus images [3], skin images [4], and pathology mainly on cancer [5]. Currently, the implementation of these technologies in the clinical process and their effect on healthcare workers are of great interest [6].

There are some studies trying to apply DL to renal pathology. While some studies have validated DL models analyzing the structures other than the glomeruli, such as the tubules, blood vessels, and interstitium [7–10], many studies have focused on the glomeruli, which present various histological findings essential for diagnosis. As a first step in the automation of this diagnostic procedure, detection of a glomerulus in a whole slide image (WSI) of renal tissue specimens has been recently attempted in many studies with the use of methods to define various features [11–24] or using convolutional neural networks (CNNs) [25], such as InceptionV3 [26], AlexNet [27], U-Net [28], R-CNN [29,30], or DeepLab V2 ResNet [31].

On the other hands, studies trying to classify pathologic findings from the glomerular images are still very few, and the pathological findings analyzed in these studies are quite limited. Barros et al. [32] constructed a model to classify proliferative lesions. Sheehan et al. [24] quantified mesangial matrix proliferation, numbers of nuclei, and capillary openness. Ginley et al. [31] also quantified nuclei, luminal space, and periodic acid-Schiff-positive component. Kannan et al. [26] and Marsh et al. [33] reported models to distinguish between sclerotic and nonsclerotic glomeruli. The pathological findings analyzed in these studies are quite limited, and do not cover the pathological findings necessary for accurate diagnosis, and there has been no study which enables the comprehensive evaluation of the essential pathological findings necessary for the diagnosis.

In this study, we focused on seven major pathological findings required for pathological diagnosis: global sclerosis, segmental sclerosis, endocapillary proliferation, mesangial matrix accumulation, mesangial cell proliferation, crescent, and basement membrane structural changes, and developed AI models to classify these findings. In addition, we examined whether our AI model can cooperate with nephrologists and improve their diagnostic performance. Although many studies have compared the performance between AI and the specialists [3,4], validation of the effect of the collaboration between AI and clinicians on the diagnostic judgment is also important and clinically relevant. Assuming a situation in which a majority decision of diagnosis is taken among specialists at a case conference, we demonstrated that the diagnostic performance was improved by adding AI model as one of the specialists.

## 2. Materials and Methods

### 2.1. Data preparation

We used WSIs of 283 renal biopsy cases that were agreed to be used for research at the Kyoto University Hospital between 2012 and 2017. The renal biopsy samples, including the transplanted allografts, were obtained by needle biopsy. Specimens that were stained by periodic acid-Schiff (PAS) and periodic acid methenamine silver (PAM) were used. Details of the scanning of the slides are provided in the Supplementary Material.

Patients provided written informed consent for the use of the specimens in this research. Moreover, we posted an announcement regarding this research study on our department website and provided information on exclusion from participation in the study. The study protocol was approved by the Ethics Committee on Human Research of the Graduate School of Medicine, Kyoto University (No. R643-2 and G562).

### 2.2. Annotation of images

Using the ImageJ software [34], two nephrologists annotated and recorded the positions and coordinates of the glomeruli in all the WSIs. Subsequently, the pathological findings in the cropped glomerular images were annotated using an original graphical user interface-based input system (Supplementary Figure S1). The following seven findings in all glomeruli were respectively evaluated as positive (+), undecidable (±), or negative (−): global sclerosis, segmental sclerosis, endocapillary proliferation, basement membrane structural changes, mesangial matrix accumulation, mesangial cell proliferation, and crescent formation (examples in Figure 1). In this study, basement membrane structural changes were defined as the presence of basement membrane thickening, spike formation, bubbling appearance, or double-contoured basement membrane. Additionally, for all glomeruli, the quality of the sample was evaluated and annotated as “artifact,” which represents glomeruli that were not suitable for evaluation of the findings, such as those collapsed by external forces, not in focus, or had dust on them (examples in Supplementary Figure S2).

**Figure 1.**
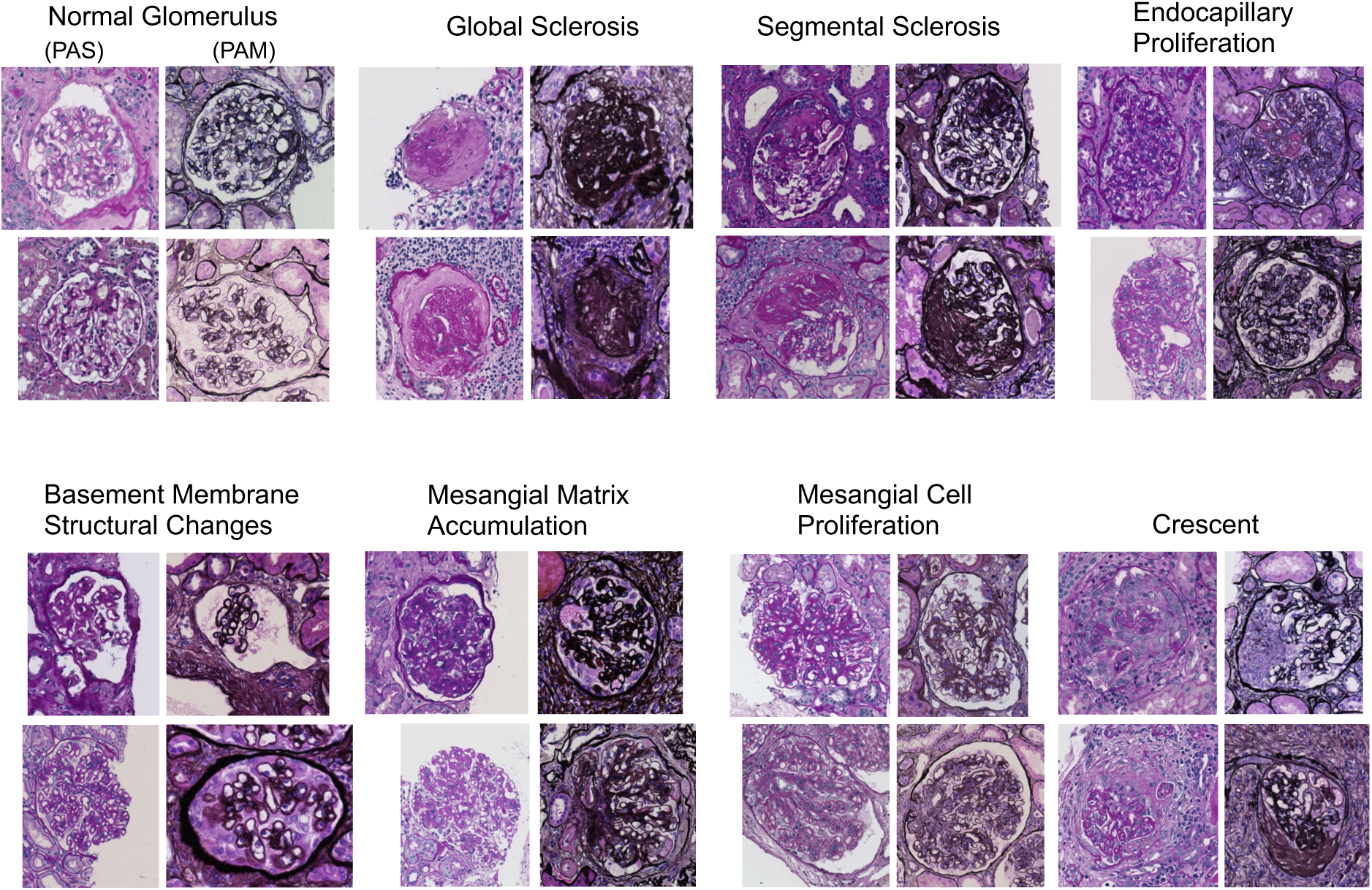
Examples of glomeruli with each pathological finding. The images show the representative glomeruli that were annotated as positive for the following findings: global sclerosis, segmental sclerosis, endocapillary proliferation, basement membrane structural changes, mesangial matrix accumulation, mesangial cell proliferation, and crescent formation. PAS, periodic acid-Schiff; PAM, periodic acid methenamine silver

In Japan, as the number of nephropathologists is still quite small, nephrologists are trained and are practicing renal pathology in most clinical situations. Thus, we asked nephrologists to annotate the datasets. The annotators were blinded to the patient information, clinical information, and diagnosis, because this study aimed at judging the findings based on the image alone.

### 2.3. Train/validation dataset

We used the images obtained between January 2012 and June 2016 as the train/validation dataset, which was used for training and parameter tuning for the AI models. The train/validation dataset was annotated by a total of 16 nephrologists. Two nephrologists were randomly allocated to each case and independently conducted the annotation (Figure 2a).

**Figure 2.**
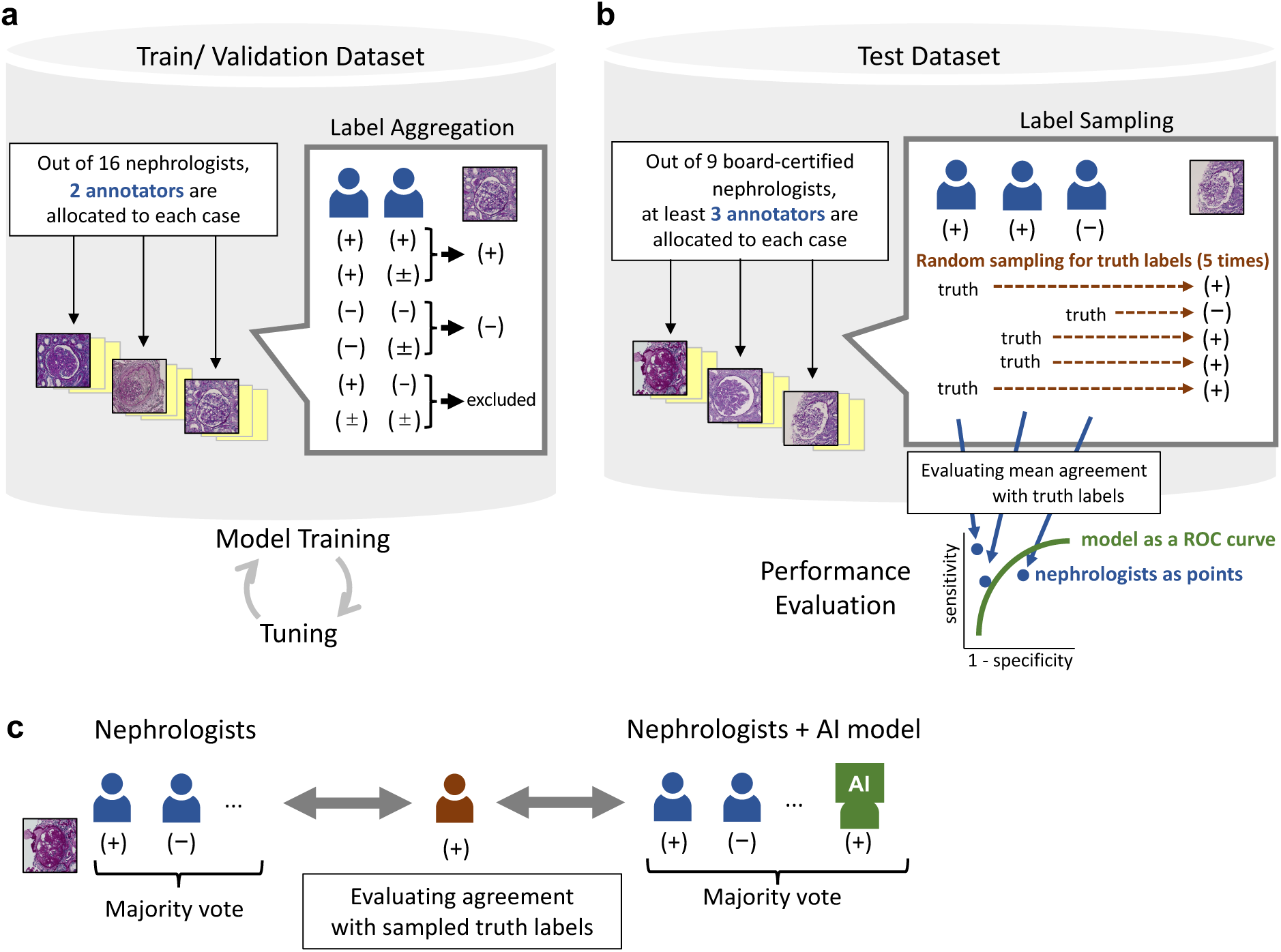
Dataset construction and framework for training and testing of the models. (a) Train/validation dataset. Out of a total of 16 nephrologists, two are assigned to each case and annotated glomerular images. For each image, two labels are aggregated to determine a truth label (label aggregation). This dataset is used for model training, tuning hyperparameters, and its validation. (b) Test dataset. Out of a total of nine board-certified nephrologists, at least three are assigned to each case and annotated glomerular images. For each image, a randomly selected label is adopted as a truth label (label sampling). This sampling process is repeated five times, and the average performance of the model is assessed in comparison with the nephrologists. (c) Nephrologist–AI collective intelligence approach by majority decision. It is examined whether adding model results to decision making can improve the performance. We compare the agreement to truth labels between majority decision among nephrologists alone and that with the AI model as a voter.

Subsequently, we performed label aggregation process in order to determine a truth label for each image (Figure 2a). For training the model, an image in the train/validation dataset was defined as positive when the respective labels annotated by the two nephrologists were (+) and (+) or (+) and (±) or as negative when the respective labels were (−) and (−) or (−) and (±).

Images that were respectively labeled as (+) and (−) or (±) and (±) were excluded from the dataset. Images with artifact labels were also excluded.

### 2.4. Development of AI models for glomerular classification

We constructed models to classify a glomerular image as positive or negative for each pathological finding in each staining (Figure 3). We performed fine-tuning with inceptionV3 [35], which is widely used for the classification problems of other medical images [3,4], with TensorFlow [36] as the backend. The models were trained and tested separately for each pathological finding. The technical details are provided in the Supplementary Material.

**Figure 3.**
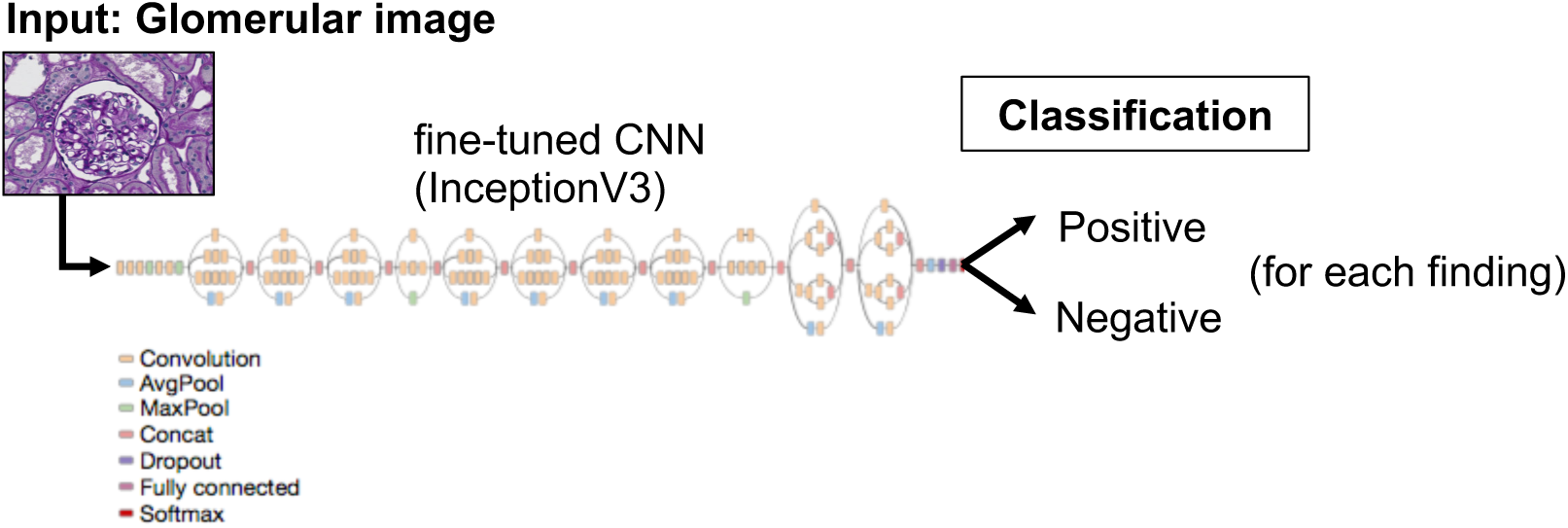
Model abstract. Classification of glomeruli by the fine-tuned InceptionV3 CNN in each finding. The model is trained for each of the seven findings of global sclerosis, segmental sclerosis, endocapillary proliferation, basement membrane structural changes, mesangial matrix accumulation, mesangial cell proliferation, and crescent formation. The outputs of the models are the degree of a positive finding when a cropped glomerular image is inputted. CNN, convolutional neural network

### 2.5. Test Dataset

We used the images obtained between July 2016 and June 2017 as the test dataset to evaluate the performances of the models (Figure 2b). The test dataset was annotated by a total of nine nephrologists, who were different from the evaluators of the train/validation dataset and had been board certified by the Japanese Society of Nephrology. At least three of the nephrologists were assigned to each case, and annotation was performed independently. Specifically for relatively rare remarks, such as crescent formation or basement membrane structural changes, cases with pathological diagnosis containing such remarks were allocated to a maximum of six nephrologists to attain enough numbers of positive labels for evaluating model performance and its comparison to nephrologists.

### 2.6. Performance evaluation of the models and nephrologists

The performance of the models was evaluated by the test dataset. The glomeruli with artifacts labels were excluded from the dataset in advance. We performed 5-time label sampling processes to determine a truth label for each image and to compare the performance between each nephrologist and the model (Figure 2b). For each image in the test dataset, an annotated label by a randomly selected nephrologist was adopted as a truth label. The annotated labels by the other nephrologists were used to evaluate their performances. Images that were sampled as truth labels of undecidable (±) were excluded from the dataset within that sampling process. This sampling process was repeated five times, and the average performance of the model was calculated. The mean performance of each nephrologist was also calculated in the five sampling results. Images that were labeled as truth by sampling were excluded from the calculation of the sampled nephrologist’s own performance.

The trained models were evaluated by area under the curve (AUC) of receiver operating characteristic (ROC) curve, sensitivity (or recall, true positive rate), and specificity (true negative rate). The performance of each annotator of the test dataset was evaluated by sensitivity and specificity.

### 2.7. Performance evaluation of the majority decision among nephrologists with the AI models

We examined whether the results of our models improved the sensitivity and specificity of the nephrologists in classifying each finding. We compared the agreement to truth labels between majority decision among nephrologists alone and that with the AI model as a voter (Figure 2c). The truth labels were sampled by the same method stated above. A nephrologist whose label was chosen as truth was excluded from the voting. In the majority decision, when the number of positive and negative judgments in an image was the same, the result was randomly decided. Statistical analyses were described in the Supplementary Material.

## 3. Results

### 3.1. Patients and annotation of images

The train/validation and test datasets included 218 and 65 cases, respectively. The demographics and pathological diagnoses of these cases are shown in Table 1. The median numbers of annotated images by one nephrologist were 1625 (532–1698 [minimum, maximum]). In the train/validation dataset, the cropped glomerular images comprised of 5571 images on PAS staining and 5876 images on PAM staining. After removing the images labeled as artifact least one annotator, 4489 images on PAS staining and 4744 images on PAM staining were used for model construction. The numbers of annotated labels in each finding are shown in Table 2. In the test dataset, the cropped glomerular images comprised 2175 images on PAS staining and 2266 images on PAM staining. After removing the images labeled as artifact by at least one annotator, 1704 images on PAS staining and 1777 images on PAM staining were used for performance evaluation. The numbers of annotated labels in each finding are shown in Table 3.

**Table 1.** Baseline case characteristics of the datasets.

|  | Train/validation dataset<br>(N = 218) | Test dataset<br>(N = 65) |
| --- | --- | --- |
| Sex |  |  |
| Male | 110 (50.5%) | 33 (50.8%) |
| Female | 108 (49.5%) | 32 (49.2%) |
| Age, years |  |  |
| Mean (SD) | 52.1 (18.7) | 50.2 (19.7) |
| Median (IQR) | 51 (36–69) | 47 (32–68) |
| Serum creatinine, mg/dL |  |  |
| Mean (SD) | 1.45 (1.25) | 1.25 (1.14) |
| Median (IQR) | 1.02 (0.73–1.64) | 0.96 (0.65–1.52) |
| Pathological diagnosis |  |  |
| IgA nephropathy | 35 (16.1%) | 17 (26.2%) |
| Lupus nephritis | 25 (11.5%) | 7 (10.8%) |
| Membranous nephropathy | 23 (10.6%) | 3 (4.6%) |
| Mesangial proliferative glomerulonephritis | 23 (10.6%) | 8 (12.3%) |
| Crescentic glomerulonephritis | 12 (5.5%) | 1 (1.5%) |
| Diabetic nephropathy | 12 (5.5%) | 3 (4.6%) |
| ANCA-associated crescentic glomerulonephritis | 11 (5.0%) | 3 (4.6%) |
| Focal segmental glomerulosclerosis | 11 (5.0%) | 2 (3.1%) |
| Minimal change nephrotic syndrome | 11 (5.0%) | 3 (4.6%) |
| Sclerosing glomerulonephritis | 11 (5.0%) | 2 (3.1%) |
| Interstitial nephritis | 9 (4.1%) | 3 (4.6%) |
| Henoch-Schönlein purpura nephritis | 8 (3.7%) | 1 (1.5%) |
| Anti-glomerular basement membrane glomerulonephritis | 4 (1.8%) | 0 (0%) |
| Endocapillary proliferative glomerulonephritis | 3 (1.4%) | 1 (1.5%) |
| Others | 20 (9.2%) | 11 (16.9%) |

ANCA, antineutrophil cytoplasmic antibody; SD, standard deviation; IQR, interquartile range

**Table 2.** Annotated labels for the train/validation dataset.

|  | Positive label |  | Negative label |  | Excluded |  |
| --- | --- | --- | --- | --- | --- | --- |
| Global sclerosis |  |  |  |  |  |  |
| PAS | 834 | (18.6%) | 3399 | (75.7%) | 256 | (5.7%) |
| PAM | 819 | (17.3%) | 3663 | (77.2%) | 262 | (5.5%) |
| Segmental sclerosis |  |  |  |  |  |  |
| PAS | 25 | (0.6%) | 4240 | (94.5%) | 224 | (5.0%) |
| PAM | 19 | (0.4%) | 4525 | (95.4%) | 200 | (4.2%) |
| Endocapillary proliferation |  |  |  |  |  |  |
| PAS | 104 | (2.3%) | 4018 | (89.5%) | 367 | (8.2%) |
| PAM | 100 | (2.1%) | 4258 | (89.8%) | 386 | (8.1%) |
| Basement membrane structural changes |  |  |  |  |  |  |
| PAS | 66 | (1.5%) | 4142 | (92.3%) | 281 | (6.3%) |
| PAM | 101 | (2.1%) | 4211 | (88.8%) | 432 | (9.1%) |
| Mesangial matrix accumulation |  |  |  |  |  |  |
| PAS | 582 | (13.0%) | 2953 | (65.8%) | 954 | (21.3%) |
| PAM | 272 | (5.7%) | 3567 | (75.2%) | 905 | (19.1%) |
| Mesangial cell proliferation |  |  |  |  |  |  |
| PAS | 304 | (6.8%) | 3426 | (76.3%) | 759 | (16.9%) |
| PAM | 59 | (1.2%) | 4219 | (88.9%) | 466 | (9.8%) |
| Crescent formation |  |  |  |  |  |  |
| PAS | 112 | (2.5%) | 4159 | (92.7%) | 218 | (4.9%) |
| PAM | 124 | (2.6%) | 4435 | (93.4%) | 185 | (3.9%) |

PAS, periodic acid-Schiff; PAM, periodic acid methenamine silver

**Table 3.** Annotated labels for the test dataset.

|  | Positive label |  | Negative label |  | Excluded |  |
| --- | --- | --- | --- | --- | --- | --- |
| Global sclerosis |  |  |  |  |  |  |
| PAS | 231.8 ± 3.3 | (13.5%) | 1459.4 ± 6.2 | (84.8%) | 12.8 ± 3.5 | (0.7%) |
| PAM | 250.2 ± 6.2 | (13.9%) | 1516.4 ± 6.2 | (84.2%) | 10.4 ± 2.3 | (0.6%) |
| Segmental sclerosis |  |  |  |  |  |  |
| PAS | 44.8 ± 5.4 | (2.6%) | 1651.8 ± 5.6 | (96.0%) | 7.4 ± 2.3 | (0.4%) |
| PAM | 35.6 ± 1.8 | (2.0%) | 1734.6 ± 3.4 | (96.3%) | 6.8 ± 2.3 | (0.4%) |
| Endocapillary proliferation |  |  |  |  |  |  |
| PAS | 96.0 ± 4.9 | (5.6%) | 1572.2 ± 5.6 | (91.4%) | 35.8 ± 4.8 | (2.1%) |
| PAM | 64.2 ± 3.3 | (3.6%) | 1672.6 ± 6.8 | (92.8%) | 40.2 ± 4.1 | (2.2%) |
| Basement membrane structural changes |  |  |  |  |  |  |
| PAS | 40.2 ± 3.8 | (2.3%) | 1649.2 ± 6.7 | (95.8%) | 14.6 ± 3.9 | (0.8%) |
| PAM | 82.0 ± 2.3 | (4.6%) | 1671.8 ± 4.0 | (92.8%) | 23.2 ± 4.1 | (1.3%) |
| Mesangial matrix accumulation |  |  |  |  |  |  |
| PAS | 847.6 ± 11.5 | (49.3%) | 825.2 ± 11.3 | (47.9%) | 31.2 ± 1.3 | (1.8%) |
| PAM | 717.0 ± 11.8 | (39.8%) | 1033.2 ± 12.7 | (57.3%) | 26.8 ± 5.4 | (1.5%) |
| Mesangial cell proliferation |  |  |  |  |  |  |
| PAS | 667.4 ± 19.9 | (38.8%) | 989.2 ± 19.3 | (57.5%) | 47.4 ± 2.3 | (2.8%) |
| PAM | 430.0 ± 12.7 | (23.9%) | 1298.8 ± 13.6 | (72.1%) | 48.2 ± 2.6 | (2.7%) |
| Crescent formation |  |  |  |  |  |  |
| PAS | 44.4 ± 6.9 | (2.6%) | 1645.6 ± 9.1 | (95.6%) | 14.0 ± 4.6 | (0.8%) |
| PAM | 39.6 ± 3.0 | (2.2%) | 1721.4 ± 3.5 | (95.9%) | 16.0 ± 1.9 | (0.9%) |

Values are expressed as mean ± standard deviation in the five sampling iterations.
PAS, periodic acid-Schiff; PAM, periodic acid methenamine silver

### 3.2. Performance of AI models for glomerular classification

The performances of the models for classification of each pathological finding on PAS and PAM staining are shown in Figures 4 and 5, respectively. The nephrologists showed high agreement for global sclerosis in both staining, and the models also showed high performance, with an AUC of 0.98. The classification examples of global sclerosis are shown in Supplementary Figure S3. In the other findings, the performance of the model ranged from an AUC of 0.59 to 0.87, with performance variation among nephrologists. For segmental sclerosis, endocapillary proliferation, membrane proliferation, and crescent formation, the nephrologists showed high specificity, but the sensitivity largely varied among them. Therefore, we evaluated the sensitivity of each model output based on a cutoff value that was the closest to the specificity of the nephrologists. The sensitivity of each model was lower than the average sensitivity of the nephrologists but exceeded the sensitivity of some nephrologists (Figures 4 and 5).

**Figure 4.**
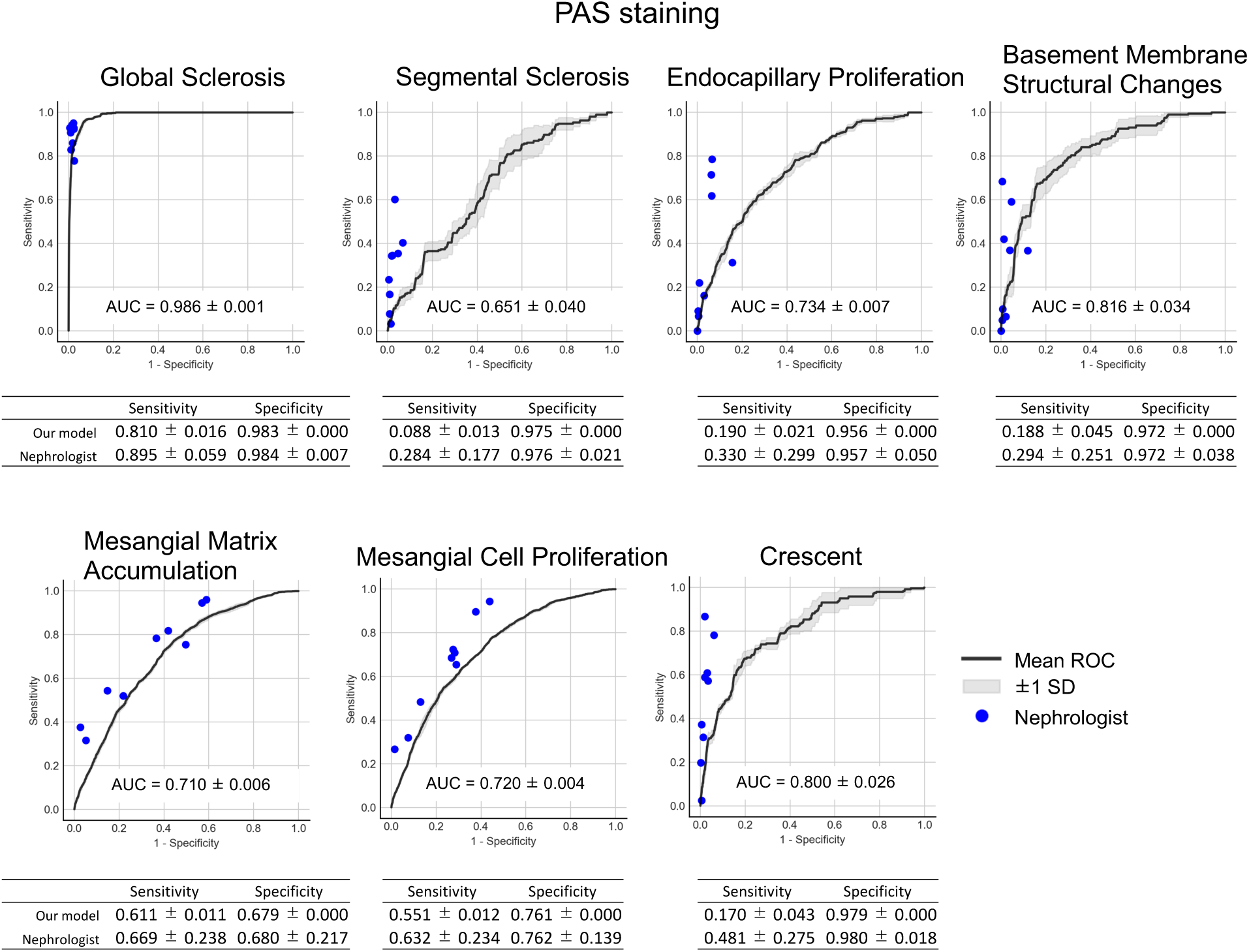
Performance of glomerular classification for seven findings on PAS staining. The performances of the models and nephrologists in the test dataset are shown as ROC curves and plotted points, respectively. Classification sensitivity and specificity are shown as mean ± SD. The cutoff points of the model outputs are determined by the nearest specificity points of the nephrologists’ mean output. PAS, periodic acid-Schiff; ROC, receiver operating characteristic; SD, standard deviation; AUC, area under the curve

**Figure 5.**
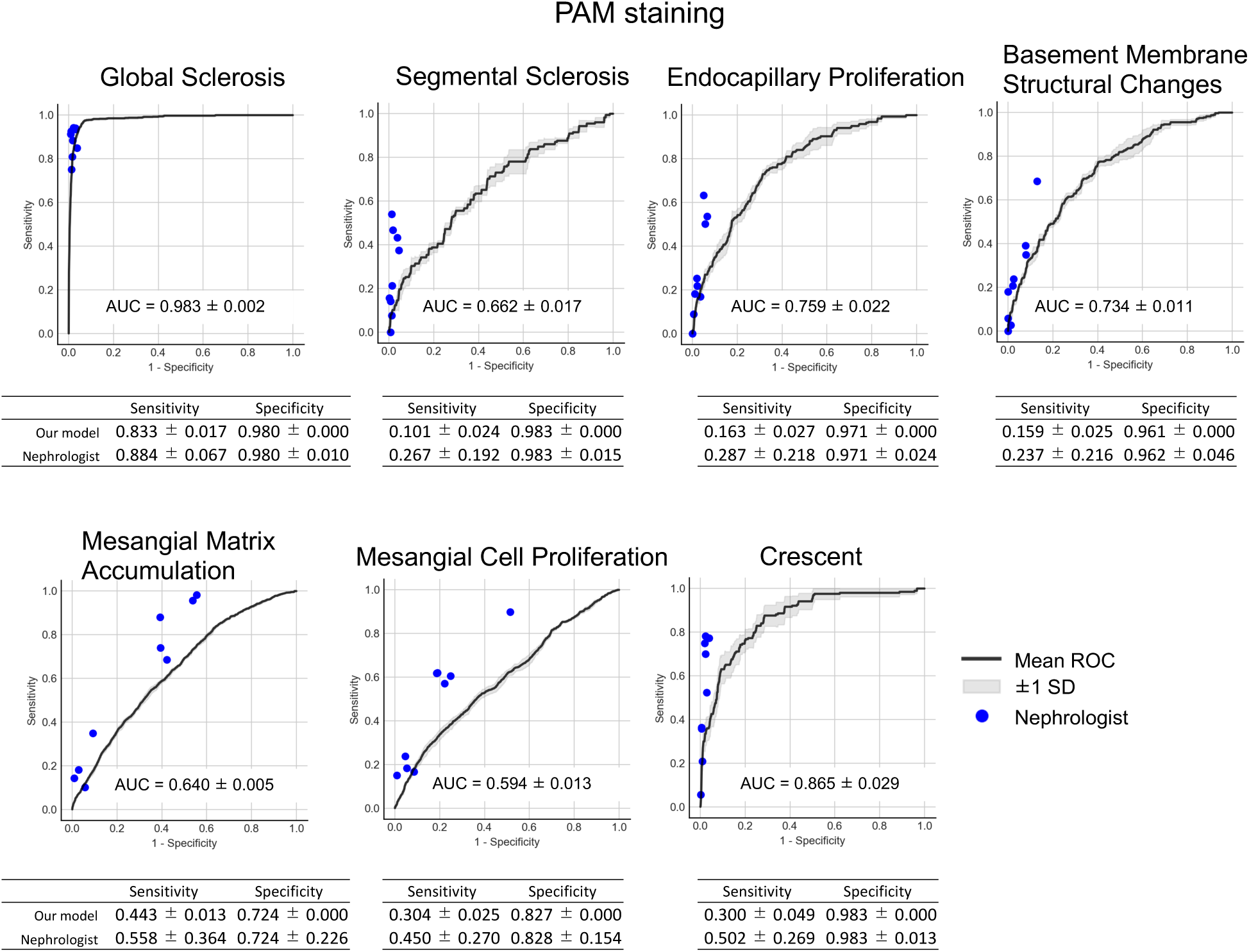
Performance of glomerular classification for seven findings on PAM staining. The performances of the models and nephrologists in the test dataset are shown as ROC curves and plotted points, respectively. Classification sensitivity and specificity are shown as mean ± SD. The cutoff points of the model outputs are determined by the nearest specificity points of the nephrologists’ mean output. PAM, periodic acid methenamine silver; ROC, receiver operating characteristic; SD, standard deviation; AUC, area under the curve

### 3.3. Performance of the majority decision among nephrologists with and without the AI

The results of the classification model were determined, based on the same cutoff value as described above for each model. The results of the majority decision showed significant improvement in specificity for all the findings in both PAS and PAM staining, and sensitivity for 9 of the 14 constructed models, when the model results were included (Table 4).

**Table 4.** Classification performance evaluation of the majority decision among nephrologists with and without

|  |  | Sensitivity |  |  | Specificity |  |  |
| --- | --- | --- | --- | --- | --- | --- | --- |
|  |  | Nephrologists | Nephrologists + AI model | p-value | Nephrologists | Nephrologists + AI model | p-value |
| Classification model |  |  |  |  |  |  |  |
| Global sclerosis |  |  |  |  |  |  |  |
|  | PAS | 0.922 ± 0.018 | 0.946 ± 0.011 | 0.038 | 0.986 ± 0.001 | 0.993 ± 0.001 | < 0.0001 |
|  | PAM | 0.907 ± 0.017 | 0.946 ± 0.023 | 0.016 | 0.985 ± 0.004 | 0.990 ± 0.002 | 0.029 |
| Segmental sclerosis |  |  |  |  |  |  |  |
|  | PAS | 0.252 ± 0.037 | 0.278 ± 0.026 | 0.23 | 0.981 ± 0.003 | 0.997 ± 0.001 | < 0.0001 |
|  | PAM | 0.241 ± 0.042 | 0.308 ± 0.061 | 0.084 | 0.987 ± 0.001 | 0.998 ± 0.001 | < 0.0001 |
| Endocapillary proliferation |  |  |  |  |  |  |  |
|  | PAS | 0.252 ± 0.022 | 0.307 ± 0.033 | 0.016 | 0.964 ± 0.003 | 0.989 ± 0.002 | < 0.0001 |
|  | PAM | 0.197 ± 0.043 | 0.286 ± 0.055 | 0.022 | 0.974 ± 0.002 | 0.993 ± 0.001 | < 0.0001 |
| Basement membrane structural changes |  |  |  |  |  |  |  |
|  | PAS | 0.269 ± 0.039 | 0.298 ± 0.031 | 0.24 | 0.985 ± 0.001 | 0.996 ± 0.001 | < 0.0001 |
|  | PAM | 0.178 ± 0.018 | 0.261 ± 0.030 | 0.0015 | 0.962 ± 0.003 | 0.992 ± 0.002 | < 0.0001 |
| Mesangial matrix accumulation |  |  |  |  |  |  |  |
|  | PAS | 0.690 ± 0.015 | 0.807 ± 0.015 | < 0.0001 | 0.650 ± 0.010 | 0.782 ± 0.009 | < 0.0001 |
|  | PAM | 0.568 ± 0.016 | 0.688 ± 0.015 | < 0.0001 | 0.705 ± 0.011 | 0.838 ± 0.010 | < 0.0001 |
| Mesangial cell proliferation |  |  |  |  |  |  |  |
|  | PAS | 0.649 ± 0.015 | 0.765 ± 0.015 | < 0.0001 | 0.755 ± 0.008 | 0.862 ± 0.008 | < 0.0001 |
|  | PAM | 0.487 ± 0.021 | 0.594 ± 0.022 | < 0.0001 | 0.826 ± 0.009 | 0.934 ± 0.002 | < 0.0001 |
| Crescent formation |  |  |  |  |  |  |  |
|  | PAS | 0.386 ± 0.043 | 0.452 ± 0.061 | 0.088 | 0.986 ± 0.003 | 0.997 ± 0.002 | 0.00050 |
|  | PAM | 0.457 ± 0.041 | 0.511 ± 0.040 | 0.071 | 0.990 ± 0.002 | 0.997 ± 0.001 | 0.0060 |

Values are expressed as mean ± standard deviation in five sampling iterations.
AI, artificial intelligence; PAS, periodic acid-Schiff; PAM, periodic acid methenamine silver

## 4. Discussion

We constructed AI models to classify several pathological findings of glomerular images. To the best of our knowledge, this is the first study to verify classification models that comprehensively included as many as seven findings essential for renal pathological diagnosis. In the classification of global sclerosis, our model showed high performance, AUC greater than 0.98, and that was also close to the performances of the nephrologists. Marsh et al. [33] reported a model to distinguish between sclerotic and nonsclerotic glomeruli on hematoxylin and eosin-stained sections of a renal graft. Their model, which used cut glomeruli beforehand, achieved a sensitivity of 0.865 and a specificity of 0.962 with the classification of glomerulosclerosis.

Although these values are not comparable directly with our study because they vary with the cutoff of the output value from the model, the discriminative ability of our model was almost the same for global sclerosis (Figures 4 and 5). There has been no previous report comparing experts and AI models in the renal pathology area. Our results showed that the current state-of-the-art AI models have almost equivalent performance to experts in classifying global sclerosis glomeruli. In the other findings, our AUC values were lower (0.6–0.8) than the results for global sclerosis. As for these findings, comparison of performance is difficult due to the difference of viewpoint from the existing research [24,32]. However, in all the findings, including crescents and membranous lesions that have not been reported for classification by AI, the present study showed performance close to nephrologists by our common method in all the findings. This result suggests the usefulness of DL in classifying many findings in renal pathology. In future studies, evaluation of the other findings, compared to global sclerosis, would need to focus on the specific parts of the glomeruli. For example, compared to an entire glomerulus, the endocapillary area, basement membrane, mesangial area, and extracapillary area are very fine structures. Moreover, there may be segmental findings that correspond to only a portion of the glomeruli. Therefore, performance may be improved by tuning, such as inputting images that are divided finely, rather than as an entire glomerulus. Some previous studies [32] showed that combination of quantification techniques may be important for findings that clinically need to be quantified, such as mesangial proliferation. In automation systems that use machine learning in other fields, performance can be improved by ensemble learning [37], which combines multiple machine learning models. In addition to the present technique, combination with these models or rule-based algorithms may further improve performance.

As an important point in our study, overall performance tended to improve when the output of the models was combined with the majority decision in nephrologists, compared to the majority decision in nephrologists alone. Notably, the current CNN can automatically extract general features from the training data [38], but it is difficult to correctly predict what greatly deviates from the data. In particular, in the pathological images with various phenotypes, compensation for situations, such as unprecedented or complicated results with pathophysiological theory or empirical knowledge, may be necessary. It is important to pay careful attention to how AI models and specialists can cooperate; however, only a few studies reported on the improvement in the prediction with the combined decision of humans and AI [39–41], and the effectiveness of a clinical decision support system that uses the AI technique has not been sufficiently verified [6]. In the research field on clinical decision support, collective intelligence approach has recently attracted attention, based on the reported higher diagnostic performance by several specialists than by a single specialist [42,43]. Although the performance of our models alone was not superior to that of the nephrologists, our results suggested that the use of the models for collective intelligence may improve the overall diagnostic performance in the clinical setting; this is a promising approach to improve the accuracy of team-based diagnosis. In future studies, it is necessary to examine how these models can actually change the decision-making or outcomes in the actual clinical setting.

This study has some limitations. A variety of annotated labels was observed among the annotators. This dataset was thought to reflect the actual variations among nephrologists, because a total of 25 annotators prepared the labels; thus, it is necessary to consider a more robust method to correct the discrepancies between evaluators. The numbers of images should be increased for the findings with the small number of positive labels, such as segmental sclerosis, although fine-tuning method had been used for training with a relatively small dataset. Our dataset was also limited to only PAS and PAM staining. Model construction and verification using a larger dataset will be required in the future.

In conclusion, we developed a classification model for seven major findings in renal pathology and demonstrated that DL method is effective for classifying these findings. We also showed that the output of the model can improve the diagnostic performance of nephrologists. The use of these models in cooperation with nephrologists, may improve the diagnostic performance for renal pathology. Further study is required to develop models that can be used in the actual clinical setting.

## Data Availability

Our data is not available because we have no permission to open our data.

## Authors’ Contributions

E.U., K.S., N.S., and Y.O. designed the study. E.U. and N.S. managed the dataset. E.U., K.S., and N.S. conducted the experiment and analyzed the data. E.U. and K.S. made the figures. R.K., Y.T., S.H., H.Y., N.Y., S.M., H.H., M.Y., and Y.O. gave administrative, technical, and material support. E.U., K.S., and N.S. drafted the manuscript. All authors revised the manuscript for important intellectual content and approved the final version of the manuscript.

### Acknowledgements

For annotation to the dataset and discussion, we thank the following nephrologists in Department of Nephrology, Graduate School of Medicine, Kyoto University: Yuki Sato, MD, PhD; Akira Ishii, MD, PhD; Keita P. Mori, MD, PhD; Naohiro Toda, MD, PhD; Keisuke Osaki, MD; Sayaka Sugioka, MD; Shinya Yamamoto, MD; Keiichi Kaneko, MD; Shunsuke Kawamura, MD; Youngna Kang, MD; Takahisa Yoshikawa, MD; Yukiko Kato, MD, PhD; Makiko Kondo, MD; Shigenori Yamamoto, MD; Yuichiro Kitai, MD; Akiko Oguchi, MD; Masahiro Takahashi, MD; Daisuke Takada, MD; Hiroyuki Arai, MD; Mitsuhiro Ichioka, MD; Koji Muro, MD; and Erina Ono, MD. We thank Kei Taneishi for his valuable technical assistance and suggestion.

This research was conducted as a joint research project and used funds from Kyoto University and Fujitsu Ltd. This work was also supported by the JSPS KAKENHI Grant Number JP18H05959. This research was partly conducted under collaborative research program with RIKEN. This research was partly supported by the Japan Agency for Medical Research and Development (AMED) under Grant Number JP18gm5010002 and by World Premier International Research Center Initiative (WPI), MEXT, Japan.

## Disclosure

E. Uchino and Y. Tamada were given a budget for a joint research project with Fujitsu Ltd. M. Yanagita received research grants from Astellas, Chugai, Daiichi Sankyo, Kyowa Hakko Kirin, Mitsubishi Tanabe Pharma Corporation, MSD, Baxter, Takeda Pharmaceutical, KISSEI PHARMACEUTICAL, Dainippon Sumitomo Pharma, TAISHO TOYAMA PHARM, and Torii. The other authors declare no conflicts of interest.

## Supplementary Material

**S1. SUPPLEMENTARY METHODS**

**S1.1. Preparation of whole slide images**

**S1.2. Technical details of the fine-tuned CNN models for glomerular classification**

**S1.3. Statistics**

**S2. SUPPLEMENTARY FIGURES**

**Supplemental Figure S1**. Graphical user interface-based annotation system for the pathological findings

**Supplemental Figure S2**. Examples of glomeruli with artifact labels

**Supplemental Figure S3**. Examples of correct and error classification of global sclerosis on PAS staining

